# Gout and coronavirus disease-19 (COVID-19): the risk of diagnosis and death in the UK Biobank

**DOI:** 10.1101/2021.09.28.21264270

**Authors:** Ruth K Topless, Angelo Gaffo, Lisa K Stamp, Philip C Robinson, Nicola Dalbeth, Tony R Merriman

## Abstract

**Background:** Data on outcomes for people with gout and COVID-19 are extremely few. Our primary objective was to assess whether gout is a risk factor for diagnosis of COVID-19 and death related to COVID-19. The secondary objectives were to test for sex- and drug-specific differences in risk.

**Methods:** We used data from the UK Biobank that included 15,560 people with gout. Multivariable-adjusted logistic regression was employed in the following analyses using a case-control study design: Analysis A, to test for association between gout and COVID-19 diagnosis (n=459,837); Analysis B, to test for association between gout and death related to COVID-19 in a case-control cohort of people who died or survived with COVID-19 (n=16,336); Analysis C, to test for association between gout and death related to COVID-19 in the entire UK Biobank cohort (n=459,837); Analysis D, to stratify by prescription of urate-lowering therapy (ULT) and colchicine on the risk of death related to COVID-19 in a subset of the UK Biobank cohort with medication data (n=341,398).

**Findings:** Gout was associated with diagnosis of COVID-19 in analysis A (OR=1.2 [1.1 ; 1.3]) but not with risk of death in the COVID-19-diagnosed group in analysis B. In analysis C gout associated with risk of death related to COVID-19 in the unadjusted model (OR=3.9 [3.3 ; 4.7]), in Model 1 adjusted for demographic factors (OR=1.8 [1.5 ; 2.1]) and in the fully adjusted Model 2 (OR=1.3 [1.1 ; 1.6]). In Analysis C risk was higher in women than men in Model 1 adjusted for demographic factors (OR=3.5 [2.4 ; 5.0] and OR=1.5 [1.2 ; 1.8], respectively) with the difference maintained after additional adjustment for eight metabolic co-morbidities (OR_Men_=1.2 [0.9 ; 1.5], OR_Women_=1.9 [1.3 ; 2.9]). There were no statistically significant differences in risk of death related to COVID-19 according to prescription of ULT or colchicine.

**Interpretation:** Gout is a risk factor for death related to COVID-19 using the UK Biobank cohort with an increased risk in women with gout that was also driven by risk factors outside metabolic co-morbidities of gout.

**Research in context:** *Evidence before this study:* There are no studies investigating the risk of COVID-19 diagnosis and risk of death with COVID-19 in people with gout.

*Added value of this study:* The findings provide evidence that gout is a risk factor for diagnosis of COVID-19 and that gout is a risk factor for death with COVID-19, independent of included co-morbidities. Women with gout are at a higher risk of death with COVID-19 than men with gout.

*Implications of the available evidence:* The new evidence demonstrate that gout is a risk factor for death from COVID-19, particularly in women. This information will inform clinical decision-making in people with gout diagnosed with COVID-19. Future research should focus on replicating these findings, including a focus on understanding key factor(s) explaining the increased risk of death with COVID-19 in women with gout.

## Background

Coronavirus disease 2019 (COVID-19) outcomes for people with inflammatory rheumatic disease (IRD) are becoming increasingly available. Within a rheumatic disease cohort of 3,729 patients the COVID-19 Global Rheumatology Alliance identified, in addition to established risk factors of older age and male sex, co-morbidities associated with death to be hypertension associated with cardiovascular disease and chronic lung disease.^1^ Despite the high prevalence of gout (e.g. 2.5% in the UK population^2^) gout is under-represented in the COVID-19 Global Rheumatology Alliance cohort,^1^ included in ‘other inflammatory arthritis’ that comprises only 2.6% of the cohort.

Our previous study^3^ in the UK Biobank cohort on outcomes for people with gout and COVID-19 comprised 2,118 COVID-19-diagnosed individuals ascertained to August 2020 – the multivariable-adjusted analysis reported no evidence for gout as a risk factor for death related to COVID-19 (OR=1.2 [0.8 ; 1.7]). The aim of the present study was to test for association of gout with diagnosis and death related to COVID-19 in a considerably larger number of individuals diagnosed with COVID-19 (n=16,898) in the UK Biobank to April 2021. We also stratified by sex and prescription of ULT and colchicine.

## Participants and methods

### Data availability

This research was conducted using the UK Biobank Resource (approval number 12611). The UK Biobank is a large resource of nearly 500,000 volunteers aged 49-86 years of age at recruitment.^4^ Recruitment began in 2006 with follow-up intended for at least 30-years. SARS-CoV-2 test information, ICD-10 hospital codes, death records and general practice prescription information were obtained via the UK Biobank data portal on the 13^th^ of September 2021. This information covered hospital diagnoses between 1991 and 7^th^ May 2021, SARS-CoV-2 tests between 16^th^ March 2020 and 6^th^ April 2021, and death records up until 23^rd^ March 2021. Participants who did not have a BMI measure, Townsend index score or smoking status were removed (n = 4,469).

### Gout and COVID-19 definitions and case-control datasets

Seventeen diseases were included for two reasons – known to be a co-morbidity of gout^5^ and / or a risk factor for death related to COVID-19.^6^ The criteria for COVID-19 diagnosis were defined as participants with 1) a positive SARS-CoV-2 test and / or 2) ICD-10 code for confirmed COVID-19 (U07.1) or probable COVID-19 (U07.2) in hospital records, or death records (Figure 1). This definition resulted in identification of 16,898 individuals who were further divided into those that died (n=1,111) based on death records and those who were known to survive (n = 15,224). 563 participants who were diagnosed after 20th February 2021 (28 days before the last recorded death) were removed from the cohort used in Analysis B (below) given the unknown outcome in these individuals. Gout was ascertained in the UK Biobank using the following criteria: self-reported gout (visits 0-2); taking allopurinol or sulphinpyrazone therapy either by self-report or from linked general practice scripts (excluding those who also had hospital diagnosed lymphoma or leukemia ICD10 C81 - C96); or hospital-diagnosed gout (ICD-10 code M10)^7^, this case definition has been validated.^7,8^ The gout cohort consisted of 15,560 cases (826 diagnosed with COVID-19). We developed four case-control datasets to test for association with the following outcomes:

**Figure 1.**
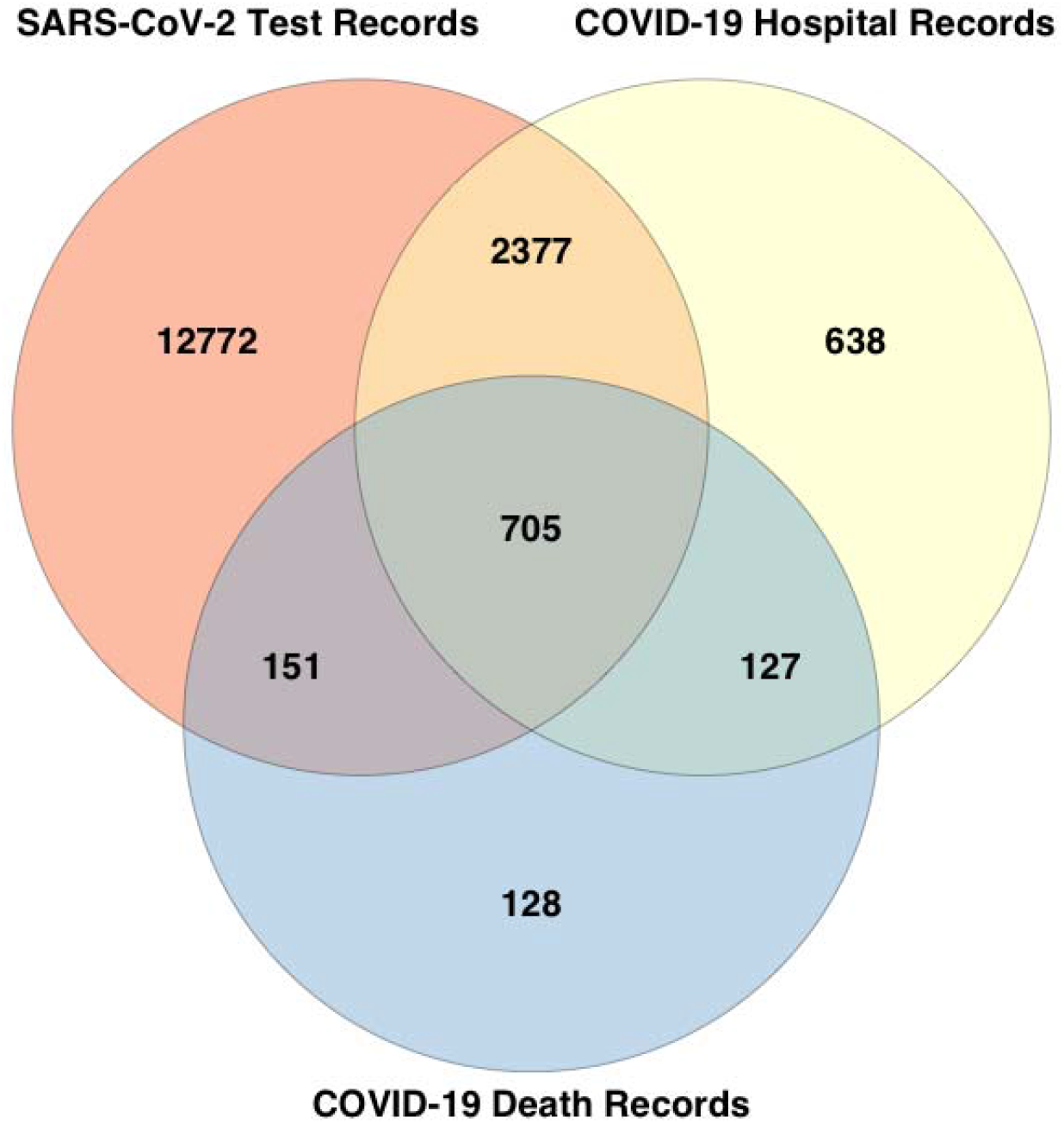
Data sources of COVID-19-diagnosed individuals. Of the 16,898 COVID-19-diagnosed individuals, 16,005 were identified from positive SARS-CoV-2 test results (12,772 unique to this group), 3,847 identified from hospital records (638 unique to this group), and 1,111 identified from death records (128 unique to this group). 563 diagnosed after 20^th^ February 2021 (28 days before the last recorded death) were removed from the cohort used in Analysis B given the unknown outcome.

Dataset A (Analysis A) to test for association with COVID-19 diagnosis in a population-based cohort. There were 16,898 people diagnosed with COVID-19 and 442,939 controls.

Dataset B (Analysis B) to test for association with death from COVID-19 in people with COVID-19. There were 1,111 people diagnosed with COVID-19 who died and 15,225 people diagnosed with COVID-19 who survived.

Dataset C (Analysis C) to test for association with death related to COVID-19 in a population-based cohort. There were 1,111 people diagnosed with COVID-19 who died and 458,726 others that included 15,787 people diagnosed with COVID-19 not known to have died.

Dataset D (Analysis D) to test for association, in the subset of the UK Biobank with requisite data, of prescription of colchicine and ULT with the risk of death related to COVID-19 for people with gout. There were 690 people diagnosed with COVID-19 who died and 340,708 others that included 12,849 people diagnosed with COVID-19 not known to have died.

### Ethnicity, age and comorbidity data

Self-reported ethnicity was grouped into White British (British, Irish, White, Any other white background), Black British (African, White and Black African, Black or Black British, Caribbean, White and Black Caribbean, Any other Black background), Asian British (Asian or Asian British, Chinese, Indian, Pakistani, Bangladeshi, White and Asian, Any other Asian background), and Other (Other ethnic group, Mixed, Any other mixed background, Do not know, Prefer not to answer). Age was calculated for 2020 from year of birth. Age groups used in the analysis were <60 years (n= 87,045), 60-69 years (n= 146,924), 70-74 years (n=107,297) and >74 years (n= 118,571). The ICD-10 hospital codes used to determine additional comorbidity status were: C00 – C96 (cancer), D80 - D89 (immunodeficiencies), E08 - E13 (diabetes mellitus), E78 (disorders of lipoprotein metabolism and other lipidemias), F01-F03 (dementia), I10 - I15 (hypertensive diseases), I60 - I69 (cerebrovascular diseases), I20 - I25 (ischemic heart diseases), I26 - I28 (pulmonary heart disease), I50 (heart failure), J44 (chronic obstructive pulmonary diseases), J45 (asthma), K70.4 / K71.1 / K72 / K91.82 (liver failure); J84 (interstitial lung disease); M19.9 (osteoarthritis) and N18 (chronic kidney disease). Only hospital records prior to a positive test for SARS-CoV-2 or a diagnosis of COVID-19 were used to determine comorbidities. This was to avoid including outcomes of COVID-19 infection in the disease associations, e.g. liver failure.

### Prescription data

General practice prescription data from 1^st^ of January 2019 through to 25^th^ July 2020, available for 341,398 participants, were used to identify people prescribed colchicine and urate-lowering therapy (allopurinol, benzbromarone, febuxostat, probenecid and sulfinpyrazone).

### Statistical analysis

All association analyses were done using R v4.0.2 in RStudio 1.2.5019. Model 1 is adjustment with age group, sex, ethnicity, Townsend deprivation index, BMI, smoking status. Model 2 is Model 1 plus adjustment by the 16 other included diseases. Analysis D is a comparison of gout with and without medical treatment (colchicine / ULT) to non-gout controls, using Model 2. Analysis of sex-specific groups was done under the same modelling. An additional model consisting of Model 1 plus eight gout-related metabolic comorbidities (hypertension, dyslipidaemia, type 2 diabetes, chronic kidney disease, obesity, coronary heart disease, cerebrovascular disease, heart failure) was used to investigate the hypothesis that women with gout are at increased risk of death related to COVID-19 owing to a metabolic co-morbidity burden, with obesity defined as BMI > 30 kg/m^2^.

A *P* < 0.05 threshold indicated nominal evidence for association. Odds ratios for risk of the various diseases use the non-disease group as comparison.

This study is reported according to the Strobe statement (http://www.strobe-statement.org).

## Results

### Participant characteristics

Table 1 summarises the characteristics of the participants according to diagnosis of COVID-19 and death with COVID-19. The proportion of people with gout who died with COVID-19 was 0.86% compared to 0.24% in the entire cohort. Similarly people who died with COVID-19 had a greater proportion of metabolic-based diseases established as co-morbid with gout,^5^ for example chronic kidney disease (19.1% vs 3.2%) and diabetes mellitus (30.7% vs 7.3%). Table 2 summarises participant characteristics according to sex. As expected men were predominant in people with gout (6.4% of the entire cohort vs 1.0% for women). However as reported in more detail elsewhere,^9^ women with gout had a higher proportion of co-morbidities than men, for example chronic kidney disease (25.5% vs 13.8%) and diabetes mellitus (26.8% vs 18.0%). In the entire cohort men have a higher proportion of these conditions (Table 2). Notably, in contrast to the over-representation of men in the entire cohort who died with COVID-19 (Table 1; 64.1% vs 35.9%), a greater proportion of women with gout died with COVID-19 than did men with gout (1.3% vs 0.8%).

**Table 1.**
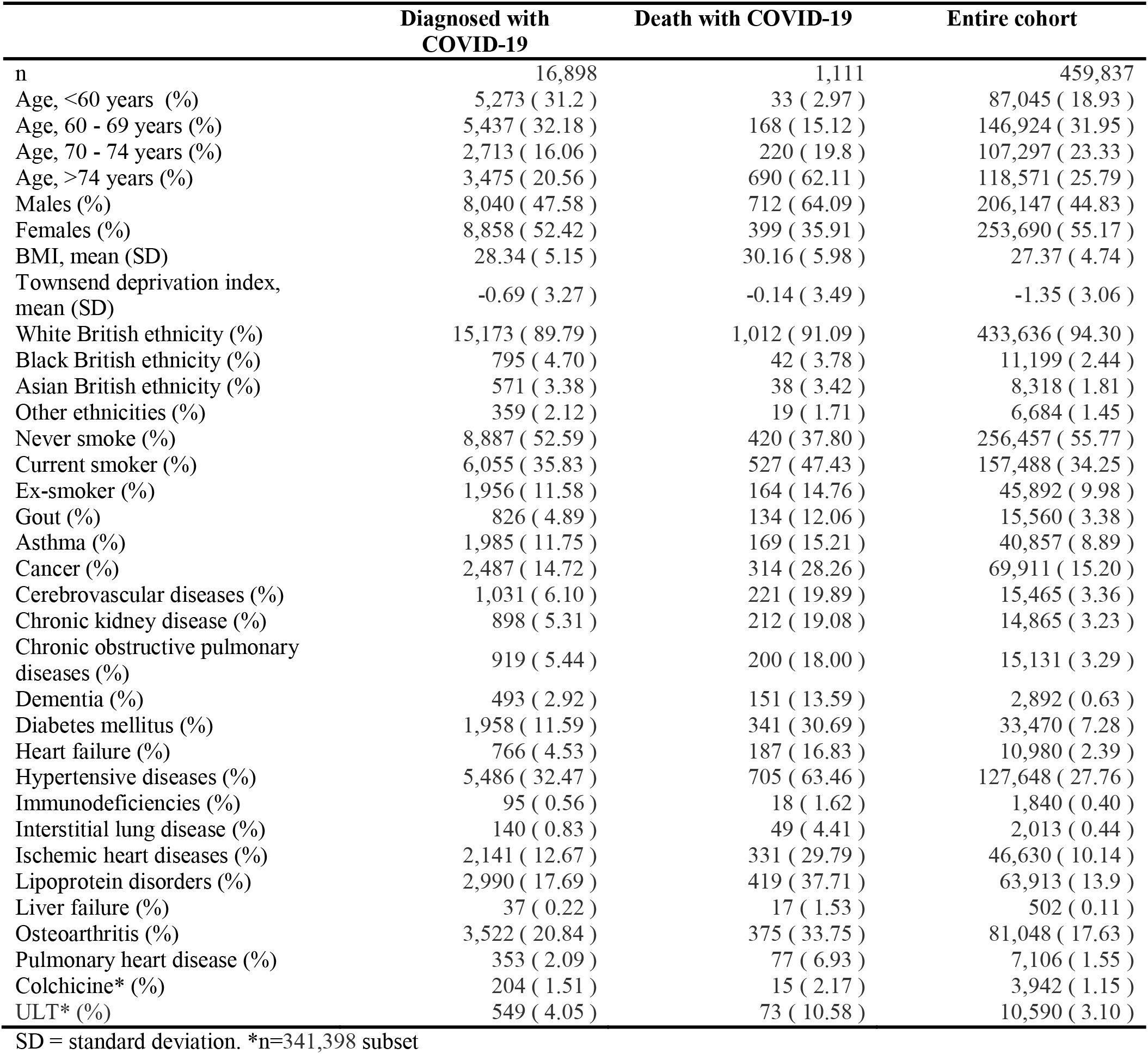
Characteristics of cohort.

|  | **Diagnosed with**<br>**COVID-19** | **Death with COVID-19** | **Entire cohort** |
| --- | --- | --- | --- |
| n | 16,898 | 1,111 | 459,837 |
| Age, <60 years (%) | 5,273 ( 31.2 ) | 33 ( 2.97 ) | 87,045 ( 18.93 ) |
| Age, 60 - 69 years (%) | 5,437 ( 32.18 ) | 168 ( 15.12 ) | 146,924 ( 31.95 ) |
| Age, 70 - 74 years (%) | 2,713 ( 16.06 ) | 220 ( 19.8 ) | 107,297 ( 23.33 ) |
| Age, >74 years (%) | 3,475 ( 20.56 ) | 690 ( 62.11 ) | 118,571 ( 25.79 ) |
| Males (%) | 8,040 ( 47.58 ) | 712 ( 64.09 ) | 206,147 ( 44.83 ) |
| Females (%) | 8,858 ( 52.42 ) | 399 ( 35.91 ) | 253,690 ( 55.17 ) |
| BMI, mean (SD) | 28.34 ( 5.15 ) | 30.16 ( 5.98 ) | 27.37 ( 4.74 ) |
| Townsend deprivation index,<br>mean (SD) | -0.69 ( 3.27 ) | -0.14 ( 3.49 ) | -1.35 ( 3.06 ) |
| White British ethnicity (%) | 15,173 ( 89.79 ) | 1,012 ( 91.09 ) | 433,636 ( 94.30 ) |
| Black British ethnicity (%) | 795 ( 4.70 ) | 42 ( 3.78 ) | 11,199 ( 2.44 ) |
| Asian British ethnicity (%) | 571 ( 3.38 ) | 38 ( 3.42 ) | 8,318 ( 1.81 ) |
| Other ethnicities (%) | 359 ( 2.12 ) | 19 ( 1.71 ) | 6,684 ( 1.45 ) |
| Never smoke (%) | 8,887 ( 52.59 ) | 420 ( 37.80 ) | 256,457 ( 55.77 ) |
| Current smoker (%) | 6,055 ( 35.83 ) | 527 ( 47.43 ) | 157,488 ( 34.25 ) |
| Ex-smoker (%) | 1,956 ( 11.58 ) | 164 ( 14.76 ) | 45,892 ( 9.98 ) |
| Gout (%) | 826 ( 4.89 ) | 134 ( 12.06 ) | 15,560 ( 3.38 ) |
| Asthma (%) | 1,985 ( 11.75 ) | 169 ( 15.21 ) | 40,857 ( 8.89 ) |
| Cancer (%) | 2,487 ( 14.72 ) | 314 ( 28.26 ) | 69,911 ( 15.20 ) |
| Cerebrovascular diseases (%) | 1,031 ( 6.10 ) | 221 ( 19.89 ) | 15,465 ( 3.36 ) |
| Chronic kidney disease (%) | 898 ( 5.31 ) | 212 ( 19.08 ) | 14,865 ( 3.23 ) |
| Chronic obstructive pulmonary<br>diseases (%) | 919 ( 5.44 ) | 200 ( 18.00 ) | 15,131 ( 3.29 ) |
| Dementia (%) | 493 ( 2.92 ) | 151 ( 13.59 ) | 2,892 ( 0.63 ) |
| Diabetes mellitus (%) | 1,958 ( 11.59 ) | 341 ( 30.69 ) | 33,470 ( 7.28 ) |
| Heart failure (%) | 766 ( 4.53 ) | 187 ( 16.83 ) | 10,980 ( 2.39 ) |
| Hypertensive diseases (%) | 5,486 ( 32.47 ) | 705 ( 63.46 ) | 127,648 ( 27.76 ) |
| Immunodeficiencies (%) | 95 ( 0.56 ) | 18 ( 1.62 ) | 1,840 ( 0.40 ) |
| Interstitial lung disease (%) | 140 ( 0.83 ) | 49 ( 4.41 ) | 2,013 ( 0.44 ) |
| Ischemic heart diseases (%) | 2,141 ( 12.67 ) | 331 ( 29.79 ) | 46,630 ( 10.14 ) |
| Lipoprotein disorders (%) | 2,990 ( 17.69 ) | 419 ( 37.71 ) | 63,913 ( 13.9 ) |
| Liver failure (%) | 37 ( 0.22 ) | 17 ( 1.53 ) | 502 ( 0.11 ) |
| Osteoarthritis (%) | 3,522 ( 20.84 ) | 375 ( 33.75 ) | 81,048 ( 17.63 ) |
| Pulmonary heart disease (%) | 353 ( 2.09 ) | 77 ( 6.93 ) | 7,106 ( 1.55 ) |
| Colchicine* (%) | 204 ( 1.51 ) | 15 ( 2.17 ) | 3,942 ( 1.15 ) |
| ULT* (%) | 549 ( 4.05 ) | 73 ( 10.58 ) | 10,590 ( 3.10 ) |

SD = standard deviation. \*n=341,398 subset

**Table 2.** Stratification of cohort by sex.

|  | All Females | All Males | Females with Gout | Males with Gout |
| --- | --- | --- | --- | --- |
| N (%) | 253,690 ( 55.20 ) | 206,147 ( 44.8 ) | 2,460 ( 0.97 ) | 13,100 ( 6.35 ) |
| COVID-19 positive | 8,858 ( 3.49 ) | 8,040 ( 3.90 ) | 165 ( 6.71 ) | 661 ( 5.05 ) |
| Death with COVID-19 | 399 ( 0.16 ) | 712 ( 0.35 ) | 33 ( 1.34 ) | 101 ( 0.77 ) |
| Age, <60 years (%) | 47,559 ( 18.75 ) | 39,486 ( 19.15 ) | 128 ( 5.20 ) | 1,277 ( 9.75 ) |
| Age, 60 - 69 years (%) | 83,269 ( 32.82 ) | 63,655 ( 30.88 ) | 510 ( 20.73 ) | 3,463 ( 26.44 ) |
| Age, 70 - 74 years (%) | 59,851 ( 23.59 ) | 47,446 ( 23.02 ) | 650 ( 26.42 ) | 3,448 ( 26.32 ) |
| Age, >74 years (%) | 63,011 ( 24.84 ) | 55,560 ( 26.95 ) | 1,172 ( 47.64 ) | 4,912 ( 37.50 ) |
| BMI, mean (SD) | 27.03 ( 5.14 ) | 27.79 ( 4.17 ) | 31.99 ( 6.39 ) | 30.08 ( 4.55 ) |
| Townsend deprivation index, mean (SD) | -1.37 ( 3.02 ) | -1.33 ( 3.11 ) | -0.73 ( 3.27 ) | -1.19 ( 3.14 ) |
| White British ethnicity (%) | 239,456 ( 94.39 ) | 194,180 ( 94.19 ) | 2,225 ( 90.45 ) | 12,415 ( 94.77 ) |
| Black British ethnicity (%) | 5,653 ( 2.23 ) | 5,546 ( 2.69 ) | 97 ( 3.94 ) | 353 ( 2.69 ) |
| Asian British ethnicity (%) | 4,889 ( 1.93 ) | 3,429 ( 1.66 ) | 71 ( 2.89 ) | 179 ( 1.37 ) |
| Other ethnicities (%) | 3,692 ( 1.46 ) | 2,992 ( 1.45 ) | 67 ( 2.72 ) | 153 ( 1.17 ) |
| Never smoke (%) | 152,649 ( 60.17 ) | 103,808 ( 50.36 ) | 1,277 ( 51.91 ) | 5,464 ( 41.71 ) |
| Current smoker (%) | 79,367 ( 31.29 ) | 78,121 ( 37.90 ) | 986 ( 40.08 ) | 6,445 ( 49.20 ) |
| Ex-smoker (%) | 21,674 ( 8.54 ) | 24,218 ( 11.75 ) | 197 ( 8.01 ) | 1,191 ( 9.09 ) |
| Asthma (%) | 24,955 ( 9.84 ) | 15,902 ( 7.71 ) | 514 ( 20.89 ) | 1,386 ( 10.58 ) |
| Cancer (%) | 36,642 ( 14.44 ) | 33,269 ( 16.14 ) | 576 ( 23.41 ) | 2,753 ( 21.02 ) |
| Cerebrovascular diseases (%) | 6,734 ( 2.65 ) | 8,731 ( 4.24 ) | 219 ( 8.90 ) | 988 ( 7.54 ) |
| Chronic kidney disease (%) | 7,525 ( 2.97 ) | 7,340 ( 3.56 ) | 627 ( 25.49 ) | 1,801 ( 13.75 ) |
| Chronic obstructive pulmonary diseases (%) | 7,353 ( 2.90 ) | 7,778 ( 3.77 ) | 262 ( 10.65 ) | 884 ( 6.75 ) |
| Dementia (%) | 1,427 ( 0.56 ) | 1,465 ( 0.71 ) | 49 ( 1.99 ) | 150 ( 1.15 ) |
| Diabetes mellitus (%) | 14,056 ( 5.54 ) | 19,414 ( 9.42 ) | 660 ( 26.83 ) | 2,354 ( 17.97 ) |
| Heart failure (%) | 4,018 ( 1.58 ) | 6,962 ( 3.38 ) | 313 ( 12.72 ) | 1,341 ( 10.24 ) |
| Hypertensive diseases (%) | 61,929 ( 24.41 ) | 65,719 ( 31.88 ) | 1,748 ( 71.06 ) | 7,657 ( 58.45 ) |
| Immunodeficiencies (%) | 983 ( 0.39 ) | 857 ( 0.42 ) | 37 ( 1.50 ) | 97 ( 0.74 ) |
| Interstitial lung disease (%) | 930 ( 0.37 ) | 1,083 ( 0.53 ) | 47 ( 1.91 ) | 144 ( 1.10 ) |
| Ischemic heart diseases (%) | 16,660 ( 6.57 ) | 29,970 ( 14.54 ) | 599 ( 24.35 ) | 3,456 ( 26.38 ) |
| Lipoprotein disorders (%) | 27,541 ( 10.86 ) | 36,372 ( 17.64 ) | 825 ( 33.54 ) | 4,169 ( 31.82 ) |
| Liver failure (%) | 188 ( 0.07 ) | 314 ( 0.15 ) | 10 ( 0.41 ) | 48 ( 0.37 ) |
| Osteoarthritis (%) | 48,027 ( 18.93 ) | 33,021 ( 16.02 ) | 1,147 ( 46.63 ) | 3,699 ( 28.24 ) |
| Pulmonary heart disease (%) | 3,431 ( 1.35 ) | 3,675 ( 1.78 ) | 147 ( 5.98 ) | 467 ( 3.56 ) |
| Colchicine* (%) | 834 ( 0.44 ) | 3,108 ( 2.06 ) | 329 ( 15.05 ) | 1,707 ( 14.77 ) |
| ULT* (%) | 1,479 ( 0.78 ) | 9,111 ( 6.04 ) | 1,479 ( 67.66 ) | 9,111 ( 78.83 ) |

SD = standard deviation. \*n=341,398 subset

### Association with diagnosis of COVID-19 (Analysis A) (Tables 3, S1, Figure 2)

Gout associated strongly with diagnosis of COVID-19 in the unadjusted analysis (OR=1.5 [1.4 ; 1.6]). In Model 1 gout associated with diagnosis (OR=1.4 [1.3 ; 1.5]). Gout remained associated with diagnosis of COVID-19 as more variables were added to the model (Model 2) – OR=1.2 [1.1 ; 1.3]. Evidenced by non-overlapping 95% CIs, women with gout were more likely to be diagnosed with COVID-19 than men in Model 2 (OR=1.5 [1.2 ; 1.7] vs 1.1 [1.0 ; 1.2]).

**Table 3.** Association analysis of gout with diagnosis and outcomes of COVID-19.

|  | Analysis A (diagnosis) |  | Analysis B (death in COVID-19 cohort) |  | Analysis C (death in entire cohort) |  |
| --- | --- | --- | --- | --- | --- | --- |
|  | OR (95% CI) | P | OR (95% CI) | P | OR (95% CI) | P |
| **Unadjusted** |  |  |  |  |  |  |
| Combined | 1.49 [ 1.39 ; 1.60 ] | 6.8x10<sup>-28</sup> | 2.98 [ 2.45 ; 3.63 ] | 1.3x10<sup>-27</sup> | 3.94 [ 3.29 ; 4.72 ] | 9.4x10<sup>-50</sup> |
| Male | 1.34 [ 1.23 ; 1.45 ] | 3.0x10<sup>-12</sup> | 1.99 [ 1.58 ; 2.50 ] | 3.5x10<sup>-09</sup> | 2.45 [ 1.98 ; 3.02 ] | 1.0x10<sup>-16</sup> |
| Female | 2.01 [ 1.71 ; 2.35 ] | 1.1x10<sup>-17</sup> | 5.74 [ 3.86 ; 8.53 ] | 6.9x10<sup>-18</sup> | 9.32 [ 6.51 ; 13.34 ] | 2.9x10<sup>-34</sup> |
| **Model 1** |  |  |  |  |  |  |
| Combined | 1.41 [ 1.31 ; 1.52 ] | 9.8x10<sup>-20</sup> | 1.44 [ 1.16 ; 1.79 ] | 1.0x10<sup>-03</sup> | 1.76 [ 1.46 ; 2.12 ] | 4.6x10<sup>-09</sup> |
| Male | 1.26 [ 1.16 ; 1.37 ] | 3.9x10<sup>-08</sup> | 1.26 [ 0.99 ; 1.62 ] | 0.063 | 1.48 [ 1.19 ; 1.84 ] | 4.0x10<sup>-04</sup> |
| Female | 1.95 [ 1.66 ; 2.30 ] | 7.5x10<sup>-16</sup> | 2.26 [ 1.45 ; 3.51 ] | 2.9x10<sup>-04</sup> | 3.47 [ 2.39 ; 5.03 ] | 6.1x10<sup>-11</sup> |
| **Model 2** |  |  |  |  |  |  |
| Combined | 1.20 [ 1.11 ; 1.30 ] | 2.5x10<sup>-06</sup> | 1.20 [ 0.95 ; 1.50 ] | 0.13 | 1.28 [ 1.05 ; 1.56 ] | 0.013 |
| Male | 1.12 [ 1.03 ; 1.22 ] | 9.7x10<sup>-03</sup> | 1.12 [ 0.86 ; 1.46 ] | 0.40 | 1.16 [ 0.93 ; 1.46 ] | 0.20 |
| Female | 1.46 [ 1.24 ; 1.73 ] | 9.1x10<sup>-06</sup> | 1.58 [ 0.98 ; 2.54 ] | 0.059 | 1.94 [ 1.30 ; 2.88 ] | 1.1x10<sup>-03</sup> |

**Figure 2.**
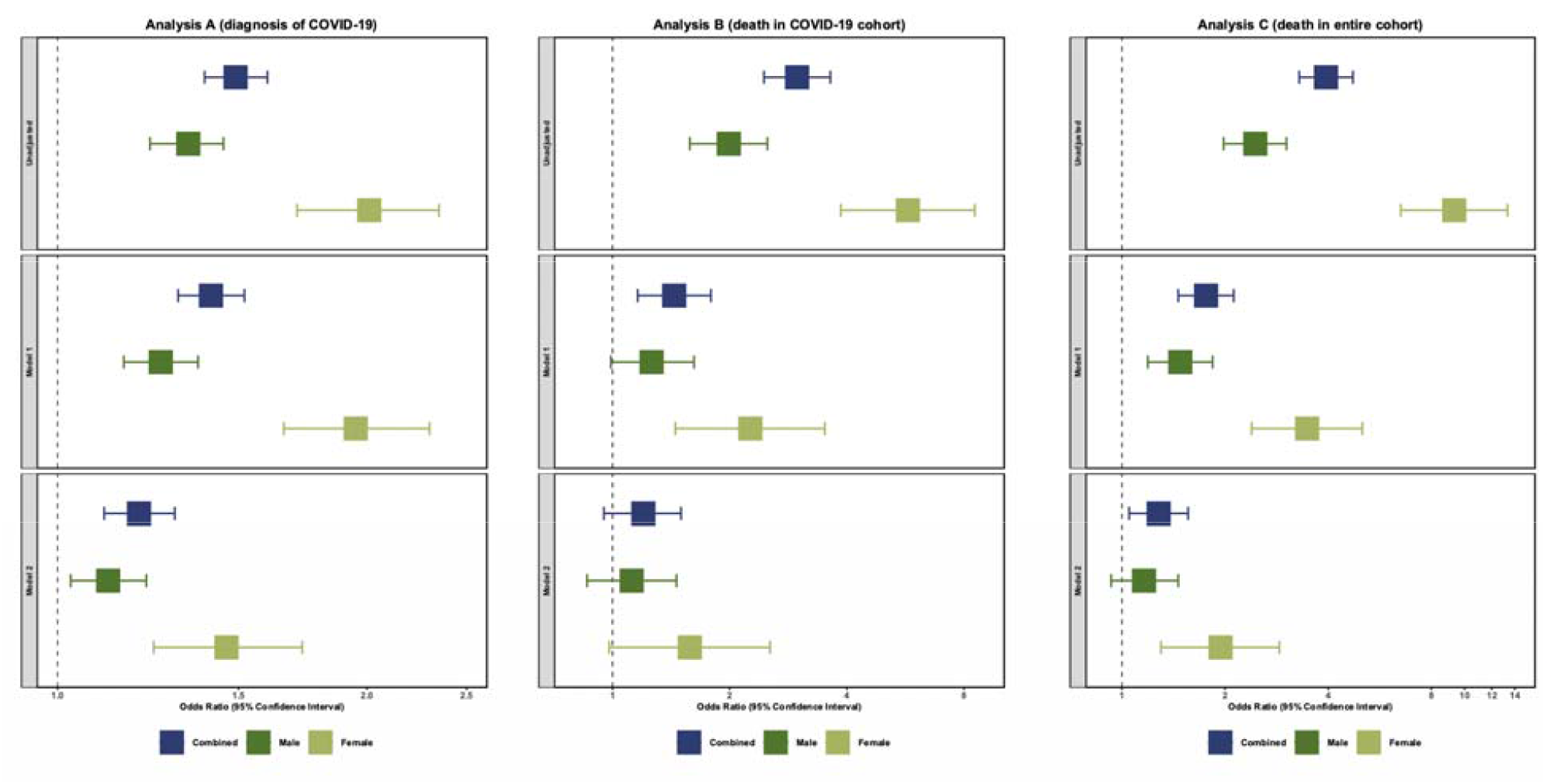
Forest plots of data presented in Table 3. Analysis A is on the left, Analysis B in the middle, Analysis C on the right. Unadjusted is top, Model 1 is middle, Model 2 is bottom.

### Association analysis with death upon diagnosis of COVID-19 (Analysis B) (Tables 3, S2, Figure 2)

In the unadjusted test of association gout associated strongly with the risk of death related to COVID-19 in Analysis B, within the cohort diagnosed with COVID-19 (OR=3.0 [2.5 ; 3.6]). Association was also evident in Model 1 (OR=1.4 [1.2 ; 1.8]). In Analysis B, Model 2 gout did not associate with the risk of death related to COVID-19 (OR=1.2 [1.0 ; 1.5]). To account for improvements in outcome over time^10-12^ we also included a quarterly (3-monthly) categorical time variable in Model 2. The categorical reference group contained diagnoses between 1^st^ January 2020 and 31^st^ March 2020 (312 positive cases including 101 deaths). After the inclusion of the categorical time variable, the association of gout and death in the COVID 19 positive cohort was statistically significant (OR = 1.3 [1.1 ; 1.7]).

### Association analysis of death with COVID-19 in the entire cohort (Analysis C) (Tables 3, S3, Figure 2)

Gout associated, with very strong statistical support, with the risk of death related to COVID-19 in the population-based unadjusted analysis (Analysis C) (OR=3.9 [3.3 ; 4.7]). Association with death was also evident in Model 1 (OR=1.8 [1.5 ; 2.1]). In Model 2 (that was additionally adjusted for the other diseases) gout associated with the risk of death (OR=1.3 [1.1 ; 1.6]).

### Sex-stratified association with death upon diagnosis of COVID-19 (Analysis C) (Table 3, Figure 2)

Women with gout were at particularly high risk of death with COVID-19 in the unadjusted Analysis C (OR=9.3 [6.5 ; 13.3]) with a 95% CI that did not overlap that of men (OR=2.5 [2.0 ; 3.0]). In Model 1 (adjusting for age, ethnicity, Townsend deprivation index^13^, BMI and smoking) the increased risk for women was OR=3.50 and OR=1.48 for men, again with non-overlapping 95% CIs (Table 3). Addition of the other 16 diseases in Model 2 resulted in increased risk of OR=1.9 [1.3 ; 2.9] for women and a statistically non-significant OR=1.2 [0.9 ; 1.5] for men, however with overlapping 95% CIs.

Women with gout have a markedly elevated risk of metabolic co-morbidities (hypertension, dyslipidaemia, type 2 diabetes, chronic kidney disease, obesity, coronary heart disease, cerebrovascular disease, heart failure),^9^ therefore to investigate the hypothesis that women with gout are at increased risk of death with COVID-19 owing to a burden of metabolic co-morbidity we added to Analysis C Model 1 only the eight aforementioned metabolic co-morbidities, with obesity defined as BMI > 30 kg/m^2^. The risk of death in women was OR=2.06 [1.39 ; 3.04], *P*=2.7×10^-4^ and men was OR=1.13 [0.91 ; 1.42], *P*=0.27 – the 95% CIs overlapped.

### Colchicine and ULT (Analysis D)

Finally, in Analysis D we stratified the UK Biobank gout cohort for prescription of each of colchicine and ULT and tested for association with death related to COVID-19 using Model 2 (using the same non-gout comparison cohort). For gout with and without colchicine the risks were 1.07 [0.60 ; 1.90] and OR=1.39 [1.08 ; 1.77], respectively, and with and without ULT the risks were OR=1.39 [1.07 ; 1.80] and OR=1.20 [0.78 ; 1.84], respectively. All 95% CIs were overlapping.

## Discussion

Gout is a very prevalent arthropathy (for example 3.9% in the US population^14^ and 14% in people of Pacific ethnicity^15^) yet has received extremely limited attention with respect to outcomes of COVID-19.^16^ Here, for the first time, we report association of gout (OR=1.3) with death related to COVID-19 in a multivariable-adjusted population-based study. Statistical support, however, is relatively weak meaning that replication in larger cohorts is necessary.

The majority of the risk of death related to COVID-19 was accounted for by eight metabolic co-morbidities in men, but not women, despite the higher risk of metabolic co-morbidity in women with gout.^9^ This is evidence for the presence of additional risk factors for death related to COVID-19 in women. What these risk factors could be was not revealed from our analysis but could be related to factors underlying the known higher risk of metabolic co-morbidity in women with gout. When we investigated the influence of colchicine we found no difference in risk of death related to COVID-19 in the gout cohort. The COLCORONA placebo-controlled randomized clinical trial provided, as yet un-replicated, evidence that colchicine reduces the risk of death or hospital admission in PCR-proven COVID-19.^17^

The highly statistically significant unadjusted risk estimate for death related to COVID-19 in the general population was OR=3.9 for gout and was particularly high for women with gout (OR=9.4). These raw estimates can be used in clinician-patient discussions regarding patient decisions to get vaccinated against SARS-Cov-2. Little is currently known about vaccine hesitancy in people with rheumatic disease. In the Italian population vaccine hesitancy is higher in people with rheumatic disease.^18,19^ The possibility of vaccine hesitancy in people with gout is of concern in the context of our data of increased risk of death related to COVID-19. While we are unable to generalize our findings to non-UK populations, in the absence of evidence to the contrary the safer approach is to assume that risks for death related to COVID-19 in other population groups are at least equivalent. For example, the prevalence of gout in the Māori population of Aotearoa New Zealand is 8% and 14% in the Pacific population, compared to 4% in the non-Māori non-Pacific population.^15^ These figures justify a targeted strategy for vaccination of Māori and Pacific people with gout in all healthcare settings.

We have identified nine limitations to our analyses. One, these analyses pertain to the population from which the UK Biobank was derived, predominantly the white European middle-aged ethnic group of the United Kingdom, and are not necessarily generalisable to other ethnic groups or other white European ethnic groups. Two, there is also no available information on recovery status so there is the possibility of additional unidentified deaths in the COVID-19 diagnosed group in Analysis B. In addition to this COVID-19 outcomes will have been influenced over the time period of this study (March 2020-April 2021) as clinical treatments evolved. We were able to account for this only in Analysis B. Limited testing outside of the hospital setting means that the full extent of SARS-CoV-2 infection is not known in this population. Thus, it is not possible to accurately compare asymptomatic or mild COVID-19 to those with more severe disease. Three, the UK Biobank dataset is also limited to those aged 49 to 86 as of 2020, a demographic with a higher case fatality ratio.^20^ This will have contributed to the inflated infection fatality ratio in the UK Biobank cohort of 6.6%, well above general population estimates of 0.5 to 1.5% (e.g. ref^21^). Therefore our findings cannot be generalised to those under 50 years of age. Four, there is the potential in Analysis B for index event (collider) bias resulting from conditioning the sample set on COVID-19 diagnosis which would serve to bias towards the null.^22^ However these limitations were addressed using the entire cohort-based approach in Analysis C. Five, the potential effect of severity in gout could not be assessed (e.g. presence / absence of tophus, flare frequency). Six, in the context of reports of a U-shaped relationship of serum urate levels and COVID-19,^23-25^ we were unable to investigate any influence of current serum urate levels on death with COVID-19 as these data are unavailable in the UK Biobank. Seven, the medicine prescription data were single script from General Practitioners only, there was no way of ascertaining adherence or whether participants were taking the medication during the COVID-19 pandemic although we attempted to account for this by only using prescription information from 2019 and 2020. Eight, we were unable to account for the effect of individual behaviour modification on outcomes. For example people with rheumatic disease in the Netherlands have been reported to be twice as likely to adhere to strict isolation measures than healthy controls^26^ and, in the UK, people with risk factors for poor outcomes from COVID-19 exhibit greater risk-mitigating behaviour.^27^ A final point is that we included co-morbidities for gout as variables in Model 2 – there is not consensus as to whether this should be done in epidemiologic studies of COVID-19 outcomes in the rheumatic diseases.^28^ Therefore we also included models with fewer adjustors (unadjusted, and adjusted by BMI and demographic factors).

In summary, we provide evidence for an increased risk of gout for death related to COVID-19. Understanding the driver(s) for the increased risk in women with gout needs to be further explored in larger datasets.

## Data Availability

The UK Biobank source data are publicly available.

## Acknowledgements

This research was conducted using the UK Biobank Resource under Application Number 12611. We sincerely thank all participants.

## Ethical approval

The UK Biobank resource was conducted under an ethical approval from the North West Multi-centre Research Ethics Committee (MREC) of the United Kingdom. The study complies with the Declaration of Helsinki and informed consent was obtained from all participants.

## Author contributions

RKT and TRM substantially contributed to study conception and design, to acquisition and analysis of data and interpretation of results. AG, LSK, PCR and ND substantially contributed to study design and interpretation of results. All authors contributed to drafting the article and critical revision and all authors approved the final version. RKT and TRM verified the data.

## Data sharing

All data utilised in this study were accessed from the publicly available UK Biobank Resource under Application Number 12611. These data cannot be shared with other investigators.

**Table S1.** Risk of diagnosis of COVID 19 (Analysis A)

|  | Diagnosed / Not<br>diagnosed<br>16,898 / 442,939 | Unadjusted |  | Model 1 \|** |  | Model 2 |  |
| --- | --- | --- | --- | --- | --- | --- | --- |
|  |  | OR [95% CI] | P | OR [95% CI] | P | OR [95% CI] | P |
| Gout | 826 / 14,734 | 1.49<br>[ 1.39 ; 1.60 ] | 6.8x10<sup>-28</sup> | 1.41<br>[ 1.31 ; 1.52 ] | 9.8x10<sup>-20</sup> | 1.20<br>[ 1.11 ; 1.30 ] | 2.5x10<sup>-06</sup> |
| Asthma | 1,985 / 38,872 | 1.38<br>[ 1.32 ; 1.45 ] | 3.6x10<sup>-40</sup> | 1.30<br>[ 1.24 ; 1.37 ] | 1.8x10<sup>-26</sup> | 1.12<br>[ 1.06 ; 1.18 ] | 1.4x10<sup>-05</sup> |
| Cancer | 2,487 / 67,424 | 0.96<br>[ 0.92 ; 1.00 ] | 0.073 | 1.13<br>[ 1.08 ; 1.18 ] | 6.5x10<sup>-08</sup> | 1.05<br>[ 1.00 ; 1.1 ] | 0.040 |
| Cerebrovascular diseases | 1,031 / 14,434 | 1.93<br>[ 1.81 ; 2.06 ] | 5.6x10<sup>-87</sup> | 2.12<br>[ 1.98 ; 2.27 ] | 5.3x10<sup>-108</sup> | 1.52<br>[ 1.42 ; 1.63 ] | 1.0x10<sup>-30</sup> |
| Chronic kidney disease | 898 / 13,967 | 1.72<br>[ 1.61 ; 1.85 ] | 1.6x10<sup>-53</sup> | 1.85<br>[ 1.72 ; 1.99 ] | 6.1x10<sup>-64</sup> | 1.25<br>[ 1.16 ; 1.35 ] | 1.0x10<sup>-08</sup> |
| Chronic obstructive<br>pulmonary diseases | 919 / 14,212 | 1.73<br>[ 1.62 ; 1.86 ] | 6.6x10<sup>-56</sup> | 1.81<br>[ 1.69 ; 1.95 ] | 1.5x10<sup>-59</sup> | 1.33<br>[ 1.23 ; 1.44 ] | 3.1x10<sup>-13</sup> |
| Dementia | 493 / 2,399 | 5.52<br>[ 5.00 ; 6.09 ] | 6.3x10<sup>-255</sup> | 6.78<br>[ 6.13 ; 7.51 ] | 1.1x10<sup>-300</sup> | 5.00<br>[ 4.5 ; 5.56 ] | 7.7x10<sup>-197</sup> |
| Diabetes mellitus | 1,958 / 31,512 | 1.71<br>[ 1.63 ; 1.80 ] | 1.5x10<sup>-104</sup> | 1.5<br>[ 1.42 ; 1.58 ] | 4.8x10<sup>-52</sup> | 1.17<br>[ 1.11 ; 1.24 ] | 2.2x10<sup>-08</sup> |
| Heart Failure | 766 / 10,214 | 2.01<br>[ 1.87 ; 2.17 ] | 2.3x10<sup>-74</sup> | 2.09<br>[ 1.93 ; 2.26 ] | 1.4x10<sup>-77</sup> | 1.41<br>[ 1.29 ; 1.54 ] | 5.6x10<sup>-15</sup> |
| Hypertensive diseases | 5,486 / 122,162 | 1.26<br>[ 1.22 ; 1.30 ] | 7.3x10<sup>-44</sup> | 1.36<br>[ 1.31 ; 1.41 ] | 1.9x10<sup>-62</sup> | 1.09<br>[ 1.04 ; 1.13 ] | 1.1x10<sup>-04</sup> |
| Immunodeficiencies | 95 / 1,745 | 1.43<br>[ 1.16 ; 1.76 ] | 7.2x10<sup>-04</sup> | 1.39<br>[ 1.13 ; 1.72 ] | 1.8x10<sup>-03</sup> | 1.10<br>[ 0.89 ; 1.35 ] | 0.40 |
| Interstitial lung disease | 140 / 1,873 | 1.97<br>[ 1.66 ; 2.34 ] | 1.5x10<sup>-14</sup> | 2.11<br>[ 1.77 ; 2.51 ] | 4.1x10<sup>-17</sup> | 1.51<br>[ 1.27 ; 1.81 ] | 5.8x10<sup>-06</sup> |
| Ischemic heart diseases | 2,141 / 44,489 | 1.30<br>[ 1.24 ; 1.36 ] | 1.8x10<sup>-28</sup> | 1.38<br>[ 1.32 ; 1.45 ] | 1.6x10<sup>-38</sup> | 1.00<br>[ 0.94 ; 1.06 ] | 0.92 |
| Lipoprotein disorders | 2,990 / 60,923 | 1.35<br>[ 1.29 ; 1.40 ] | 1.6x10<sup>-47</sup> | 1.43<br>[ 1.37 ; 1.5 ] | 3.8x10<sup>-61</sup> | 1.13<br>[ 1.07 ; 1.18 ] | 3.1x10<sup>-06</sup> |
| Liver failure | 37 / 465 | 2.09<br>[ 1.49 ; 2.92 ] | 1.7x10<sup>-05</sup> | 1.88<br>[ 1.34 ; 2.64 ] | 2.4x10<sup>-04</sup> | 1.31<br>[ 0.93 ; 1.84 ] | 0.13 |
| Osteoarthritis | 3,522 / 77,526 | 1.24<br>[ 1.19 ; 1.29 ] | 6.2x10<sup>-29</sup> | 1.36<br>[ 1.30 ; 1.41 ] | 2.2x10<sup>-50</sup> | 1.20<br>[ 1.15 ; 1.25 ] | 3.0x10<sup>-17</sup> |
| Pulmonary heart disease | 353 / 6,753 | 1.38<br>[ 1.24 ; 1.54 ] | 6.1x10<sup>-09</sup> | 1.39<br>[ 1.25 ; 1.55 ] | 3.7x10<sup>-09</sup> | 1.05<br>[ 0.94 ; 1.17 ] | 0.42 |
| Sex - Male | 8,040 / 198,107 | 1.12<br>[ 1.09 ; 1.16 ] | 2.5x10<sup>-13</sup> | 1.06<br>[ 1.02 ; 1.09 ] | 7.4x10<sup>-04</sup> | 1.04<br>[ 1.01 ; 1.07 ] | 0.015 |
| Age, <60 years | 5,273 / 81,772 | 1.00 | - | 1.00 | 0.00 | 1.00 | - |
| Age, 60 - 69 years<sup>1</sup> | 5,437 / 141,487 | 0.60<br>[ 0.57 ; 0.62 ] | 2.4x10<sup>-150</sup> | 0.60<br>[ 0.58 ; 0.62 ] | 4.8x10<sup>-143</sup> | 0.56<br>[ 0.54 ; 0.59 ] | 8.1x10<sup>-176</sup> |
| Age, 70 - 74 years<sup>1</sup> | 2,713 / 104,584 | 0.40<br>[ 0.38 ; 0.42 ] | 0.00* | 0.41<br>[ 0.39 ; 0.43 ] | 3.6x10<sup>-290</sup> | 0.35<br>[ 0.33 ; 0.37 ] | 0.00* |
| Age, >74 years<sup>1</sup> | 3,475 / 115,096 | 0.47<br>[ 0.45 ; 0.49 ] | 2.8x10<sup>-253</sup> | 0.47<br>[ 0.45 ; 0.5 ] | 4.7x10<sup>-231</sup> | 0.35<br>[ 0.33 ; 0.37 ] | 0.00* |
| White British | 15,173 / 418,463 | 1.00 | - | 1.00 | - | 1.00 | - |
| Black British ethnicity<sup>2</sup> | 795 / 10,404 | 2.11<br>[ 1.96 ; 2.27 ] | 5.7x10<sup>-87</sup> | 1.80<br>[ 1.67 ; 1.94 ] | 6.9x10<sup>-53</sup> | 1.73<br>[ 1.60 ; 1.86 ] | 1.1x10<sup>-44</sup> |
| Asian British ethnicity<sup>2</sup> | 571 / 7,747 | 2.03<br>[ 1.86 ; 2.22 ] | 4.1x10<sup>-58</sup> | 1.35<br>[ 1.24 ; 1.48 ] | 3.2x10<sup>-11</sup> | 1.35<br>[ 1.23 ; 1.48 ] | 6.7x10<sup>-11</sup> |
| Other ethnicities<sup>2</sup> | 359 / 6,325 | 1.57<br>[ 1.41 ; 1.74 ] | 3.2x10<sup>-16</sup> | 1.23<br>[ 1.10 ; 1.37 ] | 1.8x10<sup>-04</sup> | 1.23<br>[ 1.10 ; 1.37 ] | 2.3x10<sup>-04</sup> |
| Townsend deprivation<br>index | - | 1.07<br>[ 1.07 ; 1.07 ] | 2.4x10<sup>-174</sup> | 1.04<br>[ 1.04 ; 1.05 ] | 2.0x10<sup>-65</sup> | 1.04<br>[ 1.03 ; 1.04 ] | 1.2x10<sup>-42</sup> |
| BMI (kg/m<sup>2</sup>) | - | 1.04<br>[ 1.04 ; 1.04 ] | 9.5x10<sup>-162</sup> | 1.04<br>[ 1.03 ; 1.04 ] | 1.7x10<sup>-133</sup> | 1.03<br>[ 1.02 ; 1.03 ] | 4.5x10<sup>-59</sup> |
| Non-Smoker | 8,887 / 247,570 | 1.00 | - | 1.00 | - | 1.00 | - |
| Current Smoker<sup>3</sup> | 5,957 / 151,433 | 1.11<br>[ 1.08 ; 1.15 ] | 2.1x10<sup>-10</sup> | 1.18<br>[ 1.14 ; 1.22 ] | 1.1x10<sup>-21</sup> | 1.15<br>[ 1.11 ; 1.19 ] | 1.8x10<sup>-15</sup> |
| Ex-smoker<sup>3</sup> | 1,956 / 43,936 | 1.24<br>[ 1.18 ; 1.30 ] | 3.2x10<sup>-17</sup> | 1.10<br>[ 1.04 ; 1.15 ] | 4.8x10<sup>-04</sup> | 1.02<br>[ 0.96 ; 1.07 ] | 0.57 |

\* P < 1.0x10<sup>-300</sup>; \*\* The demographic and BMI estimates are derived from the model with gout as exposure.

**Table S2.** Risk of death with COVID-19 in COVID-19 positive cohort (Analysis B)

|  |  | Unadjusted |  | Model 1 |  | Model 2 |  |
| --- | --- | --- | --- | --- | --- | --- | --- |
|  | Death with<br>COVID-19 /<br>remainder<br>1,111 / 15,225 | OR [95% CI] | P | OR [95% CI] | P | OR [95% CI] | P |
| Gout | 134 / 670 | 2.98<br>[ 2.45 ; 3.63 ] | 1.3x10<sup>-27</sup> | 1.44<br>[ 1.16 ; 1.79 ] | 1.0x10<sup>-03</sup> | 1.20<br>[ 0.95 ; 1.5 ] | 0.12 |
| Asthma | 169 / 1,756 | 1.38<br>[ 1.16 ; 1.63 ] | 2.6x10<sup>-04</sup> | 1.09<br>[ 0.9 ; 1.31 ] | 0.38 | 0.88<br>[ 0.72 ; 1.07 ] | 0.19 |
| Cancer | 314 / 2,100 | 2.46<br>[ 2.14 ; 2.83 ] | 3.0x10<sup>-37</sup> | 1.31<br>[ 1.13 ; 1.52 ] | 3.8x10<sup>-04</sup> | 1.23<br>[ 1.05 ; 1.43 ] | 9.3x10<sup>-03</sup> |
| Cerebrovascular<br>diseases | 221 / 781 | 4.59<br>[ 3.9 ; 5.41 ] | 3.4x10<sup>-74</sup> | 1.98<br>[ 1.66 ; 2.36 ] | 3.9x10<sup>-14</sup> | 1.47<br>[ 1.21 ; 1.77 ] | 9.4x10<sup>-05</sup> |
| Chronic kidney<br>disease | 212 / 665 | 5.16<br>[ 4.36 ; 6.11 ] | 3.7x10<sup>-81</sup> | 2.06<br>[ 1.72 ; 2.47 ] | 9.0x10<sup>-15</sup> | 1.41<br>[ 1.15 ; 1.73 ] | 8.9x10<sup>-04</sup> |
| Chronic obstructive<br>pulmonary diseases | 200 / 695 | 4.59<br>[ 3.87 ; 5.45 ] | 2.3x10<sup>-68</sup> | 1.81<br>[ 1.49 ; 2.19 ] | 1.3x10<sup>-09</sup> | 1.46<br>[ 1.18 ; 1.81 ] | 4.7x10<sup>-04</sup> |
| Dementia | 151 / 331 | 7.08<br>[ 5.78 ; 8.67 ] | 1.9x10<sup>-79</sup> | 2.81<br>[ 2.25 ; 3.5 ] | 2.8x10<sup>-20</sup> | 2.40<br>[ 1.91 ; 3.02 ] | 9.7x10<sup>-14</sup> |
| Diabetes mellitus | 341 / 1,564 | 3.87<br>[ 3.37 ; 4.44 ] | 1.7x10<sup>-82</sup> | 1.77<br>[ 1.51 ; 2.07 ] | 1.3x10<sup>-12</sup> | 1.36<br>[ 1.14 ; 1.61 ] | 5.4x10<sup>-04</sup> |
| Heart Failure | 187 / 560 | 5.30<br>[ 4.43 ; 6.33 ] | 5.4x10<sup>-75</sup> | 1.91<br>[ 1.57 ; 2.32 ] | 6.1x10<sup>-11</sup> | 1.24<br>[ 0.99 ; 1.55 ] | 0.64 |
| Hypertensive<br>diseases | 705 / 4,632 | 3.97<br>[ 3.50 ; 4.51 ] | 1.1x10<sup>-100</sup> | 1.52<br>[ 1.32 ; 1.75 ] | 4.9x10<sup>-09</sup> | 1.13<br>[ 0.96 ; 1.33 ] | 0.14 |
| Immunodeficiencies | 18 / 75 | 3.33<br>[ 1.98 ; 5.58 ] | 5.4x10<sup>-06</sup> | 2.21<br>[ 1.26 ; 3.89 ] | 5.8x10<sup>-03</sup> | 1.65<br>[ 0.91 ; 2.98 ] | 0.097 |
| Interstitial lung<br>disease | 49 / 89 | 7.85<br>[ 5.51 ; 11.18 ] | 4.1x10<sup>-30</sup> | 4.13<br>[ 2.8 ; 6.08 ] | 7.7x10<sup>-13</sup> | 3.34<br>[ 2.23 ; 5.01 ] | 5.5x10<sup>-06</sup> |
| Ischemic heart<br>diseases | 331 / 1,753 | 3.26<br>[ 2.84 ; 3.74 ] | 2.2x10<sup>-63</sup> | 1.41<br>[ 1.22 ; 1.64 ] | 6.4x10<sup>-06</sup> | 1.02<br>[ 0.85 ; 1.21 ] | 0.84 |
| Lipoprotein<br>disorders | 419 / 2,498 | 3.08<br>[ 2.71 ; 3.51 ] | 5.4x10<sup>-66</sup> | 1.35<br>[ 1.17 ; 1.55 ] | 2.4x10<sup>-05</sup> | 0.96<br>[ 0.82 ; 1.14 ] | 0.66 |
| Liver failure | 17 / 20 | 11.81<br>[ 6.17 ; 22.62 ] | 9.2x10<sup>-14</sup> | 9.99<br>[ 4.75 ; 21 ] | 1.3x10<sup>-09</sup> | 7.18<br>[ 3.34 ; 15.43 ] | 4.3x10<sup>-07</sup> |
| Osteoarthritis | 375 / 3,056 | 2.03<br>[ 1.78 ; 2.31 ] | 2.3x10<sup>-26</sup> | 1.05<br>[ 0.91 ; 1.21 ] | 0.49 | 0.87<br>[ 0.75 ; 1.01 ] | 0.072 |
| Pulmonary heart<br>disease | 77 / 265 | 4.2<br>[ 3.24 ; 5.46 ] | 5.0x10<sup>-27</sup> | 2.06<br>[ 1.55 ; 2.74 ] | 7.0x10<sup>-07</sup> | 1.51<br>[ 1.11 ; 2.06 ] | 7.9x10<sup>-03</sup> |
| Sex - Male | 712 / 7,083 | 2.05<br>[ 1.81 ; 2.33 ] | 1.0x10<sup>-28</sup> | 1.74<br>[ 1.52 ; 2.00 ] | 1.4x10<sup>-15</sup> | 1.65<br>[ 1.43 ; 1.91 ] | 4.8x10<sup>-12</sup> |
| Age, <60 years | 33 / 5,014 | 1.00 | - | 1.00 | - | 1.00 | - |
| Age, 60 - 69 years<sup>1</sup> | 168 / 5,097 | 5.01<br>[ 3.44 ; 7.29 ] | 3.9x10<sup>-17</sup> | 4.86<br>[ 3.34 ; 7.09 ] | 1.8x10<sup>-16</sup> | 4.30<br>[ 2.94 ; 6.28 ] | 4.4x10<sup>-14</sup> |
| Age, 70 - 74 years<sup>1</sup> | 220 / 2,407 | 13.89<br>[ 9.60 ; 20.09 ] | 2.3x10<sup>-44</sup> | 13.71<br>[ 9.45 ; 19.89 ] | 3.2x10<sup>-43</sup> | 10.29<br>[ 7.04 ; 15.03 ] | 2.0x10<sup>-33</sup> |
| Age, >74 years<sup>1</sup> | 690 / 2,707 | 38.73<br>[ 27.23 ; 55.09 ] | 5.4x10<sup>-92</sup> | 38.76<br>[ 27.13 ; 55.38 ] | 9.5x10<sup>-90</sup> | 23.99<br>[ 16.59 ; 34.69 ] | 5.7x10<sup>-64</sup> |
| White British | 1,012 / 13,656 | 1.00 | - | 1.00 | - | 1.00 | - |
| Black British<br>ethnicity<sup>2</sup> | 42 / 719 | 0.99<br>[ 0.71 ; 1.38 ] | 0.14 | 1.36<br>[ 0.96 ; 1.92 ] | 0.085 | 1.37<br>[ 0.96 ; 1.95 ] | 0.085 |
| Asian British<br>ethnicity<sup>2</sup> | 38 / 519 | 0.79<br>[ 0.57 ; 1.08 ] | 0.94 | 1.46<br>[ 1.01 ; 2.13 ] | 0.046 | 1.42<br>[ 0.97 ; 2.09 ] | 0.071 |
| Other ethnicities<sup>2</sup> | 19 / 331 | 0.77<br>[ 0.49 ; 1.24 ] | 0.28 | 1.10<br>[ 0.67 ; 1.83 ] | 0.70 | 1.05<br>[ 0.62 ; 1.76 ] | 0.87 |
| Townsend<br>deprivation index | - | 1.05<br>[ 1.04 ; 1.07 ] | 8.5x10<sup>-09</sup> | 1.05<br>[ 1.03 ; 1.07 ] | 2.8x10<sup>-07</sup> | 1.04<br>[ 1.02 ; 1.06 ] | 1.4x10<sup>-04</sup> |
| BMI (kg/m<sup>2</sup>) | - | 1.07<br>[ 1.05 ; 1.08 ] | 3.9x10<sup>-33</sup> | 1.06<br>[ 1.05 ; 1.08 ] | 7.8x10<sup>-23</sup> | 1.05<br>[ 1.04 ; 1.06 ] | 2.9x10<sup>-13</sup> |
| Non-Smoker | 420 / 8,146 | 1.00 | - | 1.00 | - | 1.00 | - |
| Current Smoker<sup>3</sup> | 527 / 5,344 | 1.91<br>[ 1.67 ; 2.18 ] | 1.0x10<sup>-21</sup> | 1.17<br>[ 1.01 ; 1.35 ] | 0.036 | 1.10<br>[ 0.95 ; 1.28 ] | 0.21 |
| Ex-smoker<sup>3</sup> | 164 / 1,735 | 1.83<br>[ 1.52 ; 2.21 ] | 2.5x10<sup>-10</sup> | 1.82<br>[ 1.48 ; 2.23 ] | 1.1x10<sup>-08</sup> | 1.51<br>[ 1.21 ; 1.88 ] | 2.3x10<sup>-04</sup> |

\* P < 1.0x10<sup>-300</sup>; \*\* The demographic and BMI estimates are derived from the model with gout as exposure.

**Table S3.**
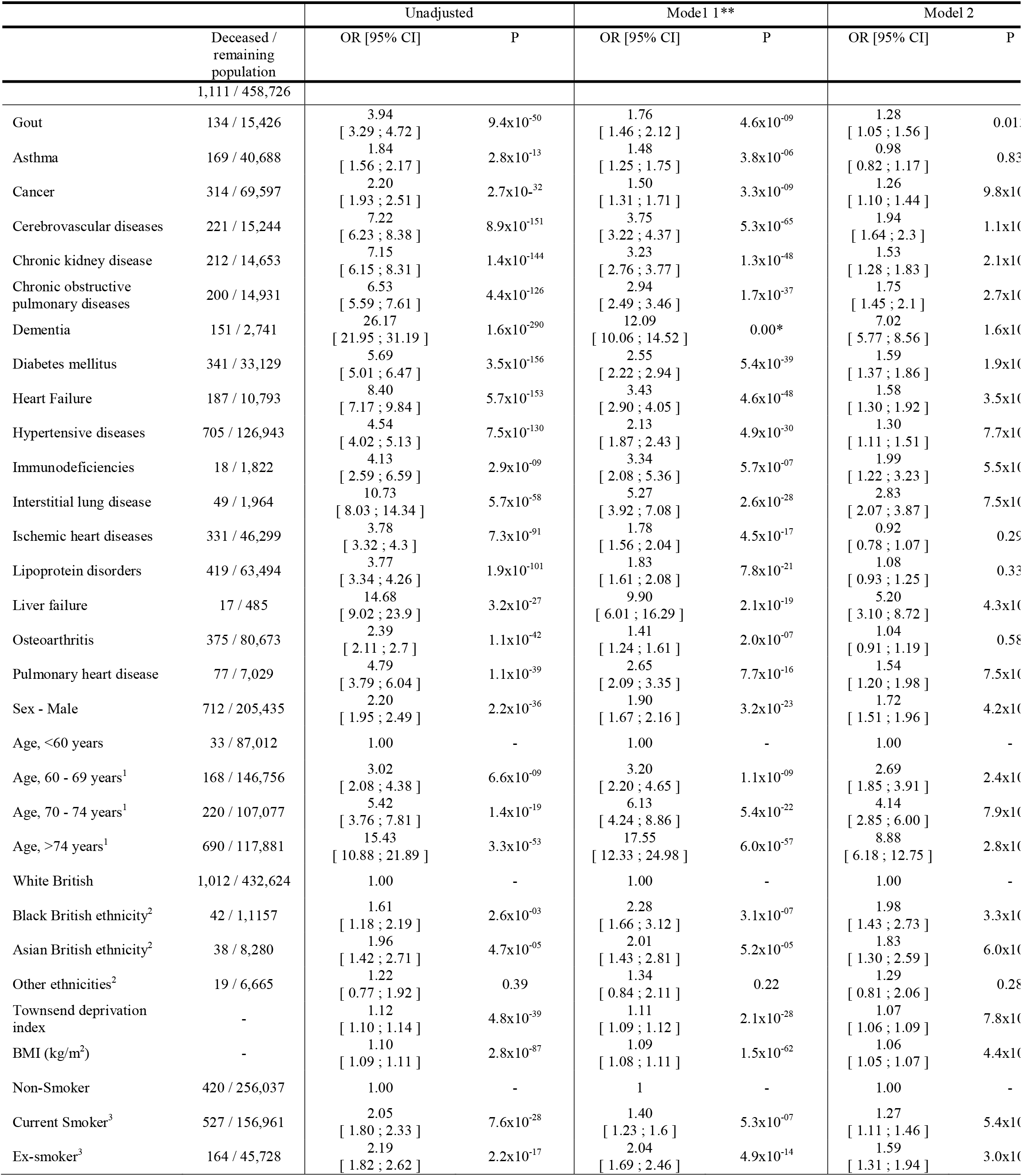
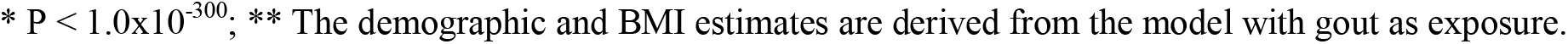
Risk of death with COVID-19 in total cohort (Analysis C)

